# Risk Assessment of Novel Coronavirus COVID-19 Outbreaks Outside China

**DOI:** 10.1101/2020.02.04.20020503

**Authors:** Péter Boldog, Tamás Tekeli, Zsolt Vizi, Attila Dénes, Ferenc A. Bartha, Gergely Röst

**Author notes:** (P.B.); (T.T.); (Z.V.); (F.A.B.); (G.R.).

## Abstract

We developed a computational tool to assess the risks of novel coronavirus outbreaks outside of China. We estimate the dependence of the risk of a major outbreak in a country from imported cases on key parameters such as: (i) the evolution of the cumulative number of cases in mainland China outside the closed areas; (ii) the connectivity of the destination country with China, including baseline travel frequencies, the effect of travel restrictions, and the efficacy of entry screening at destination; and (iii) the efficacy of control measures in the destination country (expressed by the local reproduction number *R*_loc_). We found that in countries with low connectivity to China but with relatively high *R*_loc_, the most beneficial control measure to reduce the risk of outbreaks is a further reduction in their importation number either by entry screening or travel restrictions. Countries with high connectivity but low *R*_loc_ benefit the most from policies that further reduce *R*_loc_. Countries in the middle should consider a combination of such policies. Risk assessments were illustrated for selected groups of countries from America, Asia, and Europe. We investigated how their risks depend on those parameters, and how the risk is increasing in time as the number of cases in China is growing.

## 1. Introduction

A cluster of pneumonia cases in Wuhan, China, was reported to the World Health Organization (WHO) on 31 December 2019. The cause of the pneumonia cases was identified as a novel betacoronavirus, the 2019 novel coronavirus (2019-nCoV, recently renamed as SARS-CoV-2, the cause of coronavirus disease COVID-19). The first patient showing symptoms was recorded by Chinese authorities on 8 December 2019 [1]. On 9 January 2020, WHO confirmed that a novel coronavirus had been isolated from one of the hospitalized persons [2], and the first death case was reported on the same day. The first case outside China was witnessed on 13 January in Thailand [3], and in the following days, several other countries also reported 2019-nCoV cases [4]. The first confirmed cases in China, but outside Hubei province, were reported on 19 January. [4]. As of 1 February, there were 14,628 confirmed cases worldwide (out of which 14,451 happened in China) with 305 total deaths [5].

Since no specific antiviral agent is available for treatment of this infection, and there is no vaccine [6], the control measures, introduced both in China and other countries, aimed to prevent the transmission. A metropolitan-wide quarantine of Wuhan and nearby cities was introduced on 23–24 January [7]. Several airports and train stations have started temperature screening measures to identify people with fevers [8]. All public transportation was suspended in Wuhan from 10 a.m., 23 January, including all outbound trains and flights, and all bus, metro and ferry lines; additionally, all outbound trains and flights were halted [9]. Construction of a specialist emergency hospital was started in Wuhan [10], and nearly 6000 medical workers were sent to Wuhan from across China [11]. Beijing also announced the suspension of all inter-provincial bus and train services; several touristic attractions, including the Forbidden City and Shanghai Disneyland were closed [9]. Other countries also introduced control measures, including screening passengers arriving from China and closing their borders [12]. Several airlines, including British Airways and Lufthansa, canceled all flights to and from mainland China [9].

The potential dangers of 2019-nCoV have prompted a number of studies on its epidemiological characteristics. The 2018 travel data from the International Air Transport Association (IATA) were used to identify the countries and their infectious disease vulnerability indexes (IDVIs) [13], which received substantial travel inflow from Wuhan Tianhe International Airport [14]. The IDVI has a range of 0–1, with a higher score implying lower vulnerability. The top destinations, Bangkok, Hong Kong, Tokyo and Taipei, all have an IDVI above 0.65.

It is essential to estimate the number of infections (including those that have not been diagnosed), to be able to analyze the spread of the disease. To that end, data on exported infections and individual-based mobility models were used by several researchers, obtaining comparable numbers. For 17 January 2020, preliminary estimates were given for various scenarios in the range 350–8400 by Chinazzi et al. for the total number of infections up to that date [15]. Imai et al. [16] also estimated the total number of infections in China and warned that the number is likely to substantially exceed that of the officially confirmed cases (see also [17]). They reported an estimate of 4000 infections (range: 1000–9700) by 18 January 2020. Nishiura et al. calculated 5502 (range: 3027–9057) infections by 24 January 2020 [18].

To better assess the epidemic risk of 2019-nCoV, among the key parameters to be approximated are the basic reproduction number *R*_0_ and the incubation period. We summarize previous efforts made toward those ends in Table 1, and present a short summary below.

**Table 1.**
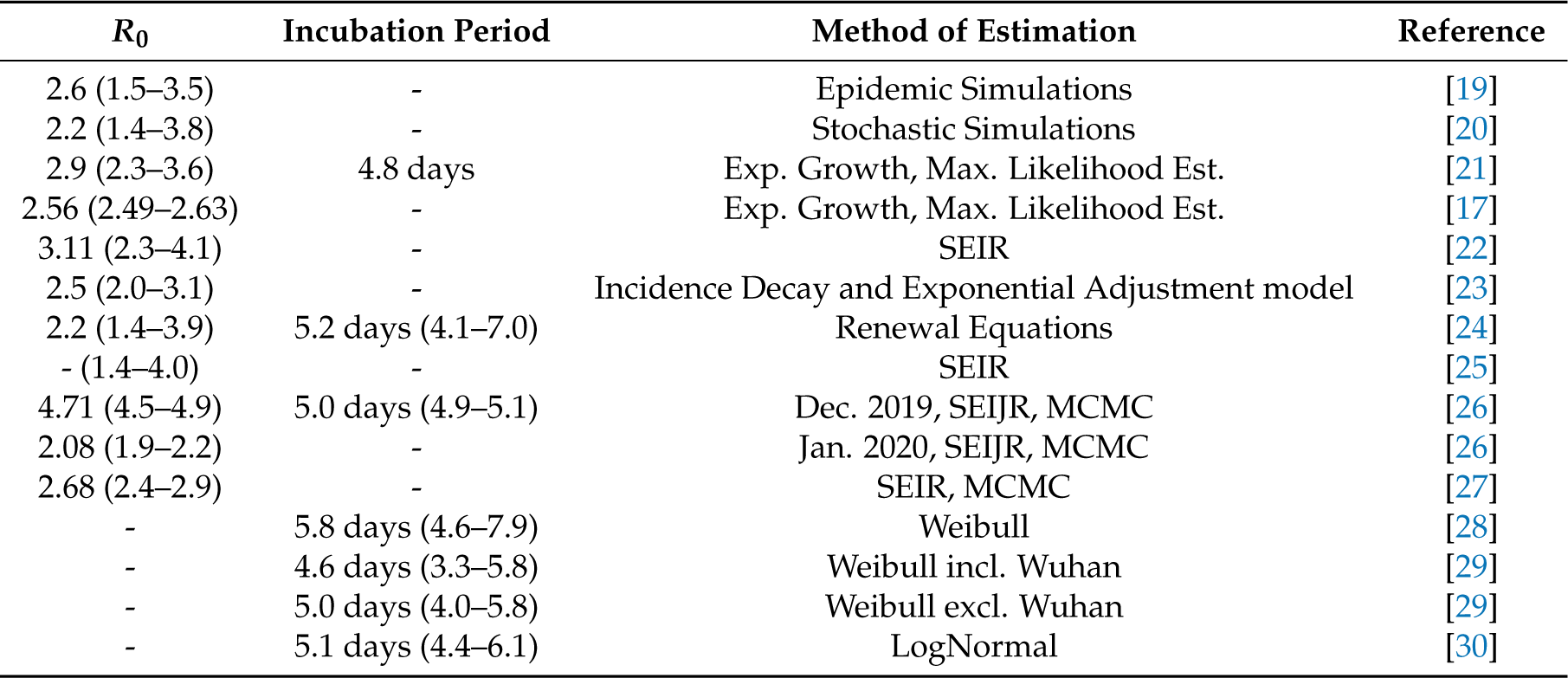
Published estimates of the key epidemiological parameters of 2019-nCoV. Uncertainty range is given where provided.

The majority of the estimates for *R*_0_ range between 2 and 3. Obtaining these was done by modeling epidemic trajectories and comparing them to the results of [16] as a baseline [19,20], using a negative binomial distribution to generate secondary infections. Liu et al. utilized the exponential growth and maximum likelihood estimation methods and found that the 2019-nCoV may have a higher pandemic risk than SARS-CoV in 2003 [21].

Read et al. based their estimates on data from Wuhan exclusively (available up to 22 January 2020) and a deterministic SEIR model [22]. The choice of this date is motivated by the actions of authorities, that is the substantial travel limitations the next day. Li et al. used solely the patient data with illness onset between 10 December 2019 and 4 January 2020 [24]. The Centre for the Mathematical Modelling of Infectious Diseases at the London School of Hygiene and Tropical Medicine have analyzed 2019-nCoV using SEIR and multiple data series [25]. Shen et al. used a SEIJR model (where J denotes the compartment of diagnosed and isolated individuals) and Markov chain Monte Carlo (MCMC) simulations [26] similarly to [27]. An alternative approach was presented by Majumder and Mandl [23] as they obtained their estimate based on the cumulative epidemic curve and the incidence decay and exponential adjustment (IDEA) model [31].

The incubation period was estimated to be in between 4.6 and 5.8 days by various studies. The first calculations used data up to 23 January [21]. Weibull distribution was identified as the best-fit model by several researches when comparing LogNormal, Gamma, and Weibull fits. Backer et al. used newly available patient data with known travel history and identified the Weibull distribution as the one with the best LOO (Leave-One-Out) score [28]; Linton et al. gave estimates for with and without Wuhan residents using their statistical model with, again, the Weibull distribution scoring the best AIC (Akaike information criterion) [29]. The Johns Hopkins University Infectious Disease Dynamics Group has been collecting substantial data on exposure and symptom onset for 2019-nCoV cases. They recommend using their LogNormal estimate [30], which gives a 5.1 day incubation period.

In this study we combine case estimates, epidemiological characteristics of the disease, international mobility patterns, control efforts, and secondary case distributions to assess the risks of major outbreaks from imported cases outside China.

## 2. Materials and Methods

### 2.1. Model Ingredients

Our method has three main components:

i. We estimate the cumulative number of cases in China outside Hubei province after 23 January, using a time-dependent compartmental model of the transmission dynamics.
ii. We use that number as an input to the global transportation network to generate probability distributions of the number of infected travellers arriving at destinations outside China.
iii. In a destination country, we use a Galton–Watson branching process to model the initial spread of the virus. We calculate the extinction probability of each branch initiated by a single imported case, obtaining the probability of a major outbreak as the probability that at least one branch will not go extinct.

### 2.2. Epidemic Size in China Outside the Closed Areas of Hubei

The starting point of our transmission model is 23 January, when major cities in Hubei province were closed [7]. From this point forward, we run a time dependent *SE*_*n*_ *I*_*m*_*R* model in China outside Hubei, which was calibrated to be consistent with the estimated case numbers outside Hubei until 31 January. We impose time dependence in the transmission parameter due to the control measures progressively implemented by Chinese authorities on and after 23 January. With our baseline *R*_0_ = 2.6, disease control is achieved when more than 61.5% of potential transmissions are prevented. We introduce a key parameter *t*_∗_ to denote the future time when control measures reach their full potential. For this study we assume it to be in the range of 20–50 days after 23 January. Using our transmission model, we calculate the total cumulative number of cases (epidemic final size) outside Hubei, for each *t*_∗_ in the given range. This also gives an upper bound for the increasing cumulative number of cases *C* = *C*(*t*).

### 2.3. Connectivity and Case Exportation

The output *C* of the transmission model is used as the pool of potential travellers to abroad, and fed into the online platform EpiRisk [32]. This way, we evaluated the probability that a single infected individual is traveling from the index areas (in our case Chinese provinces other than Hubei) to a specific destination. Using a ten day interval for potential travel after exposure (just as in [15]), one can find from EpiRisk that in the January–February periods, assuming usual travel volumes, there is a 1/554 probability that a single case will travel abroad and cause an exported case outside China. The dataset for relative importation risks of countries is available as well; thus, one can obtain the probability of an exported case appearing in a specific country. This probability is denoted by *θ*_0_, and we call it the baseline connectivity of that country with China. The baseline connectivity can be affected by other factors, such as the reduction in travel volume between the index and destination areas, exit screening in China, and the efficacy of entry screening at the destination country. Hence, we have a compound parameter, the actual connectivity *θ*, which expresses the probability that a case in China outside Hubei will be eventually mixed into the population of the destination country. For example, the relative risk of Japan is 0.13343, meaning that 13.343% of all exportations are expected to appear in Japan. Thus, under normal circumstances, the probability that a case from China eventually ends up in Japan is 0.13343/554 = 2.41 × 10^−4^ during the January–February period [32]. Assuming a 20% reduction in travel volume between China and Japan, this baseline connectivity is reduced to a connectivity 0.8 × 2.41 × 10^−4^= 1.928 × 10^−4^. Additionally, assuming a 40% efficacy on entry screening [33], there is a 0.6 probability that an arriving case passes the screening, and the connectivity parameter is further reduced to 0.6 × 1.928 × 10^−4^ = 1.16 × 10^−4^. If we assume interventions at the originating area, for example, exit screening with 25% efficacy, then our actual connectivity parameter is *θ* = 0.75 × 1.16 × 10^−4^ = 8.7 × 10^−5^, which represents the probability that a case in China will eventually mix into the population in Japan. Assuming independence, this *θ*, together with the cumulative cases *C*, generates a binomial distribution of importations that enter the population of a given country.

### 2.4. Probability of a Major Outbreak in a Country by Imported Cases

Each imported case that passes the entry screening and mixes into the local population can potentially start an outbreak, which we model by a Galton–Watson branching process with negative binomial offspring distribution with dispersion parameter *k* = 0.64 [19,20] and expectation *R*_loc_, where *R*_loc_ is the local reproduction number of the infection in a given country. Each branch has extinction probability *z*, which is the unique solution of the equation *z* = *g*(*z*) on the interval (0, 1), where *g* is the generating function of the offspring distribution (see [34]). The process dies out if all the branches die out; thus, we estimate the risk of a major local outbreak from importation as 1 − *z*^*i*^, where *i* cases were imported.

### 2.5. Dependence of the Risk of Major Outbreaks on Key Parameters

The number of imported cases *i* is given by a random variable *X*, where *X ∼* Binom(*C, θ*). The outbreak risk in a country *x* is then estimated as Risk_*x*_ = *E*[1 − *z*^*X*^], where *E* is the expectation of the outbreak probabilities; thus, we consider a probability distribution of branching processes. This way Risk_*x*_ = Risk(*C, θ, R*_loc_), which means that the risk depends on the efficacy of Chinese control measures that influence the cumulative case number *C*, the connectivity between the index and destination areas *θ*, and the local reproduction number *R*_loc_. The main question we aim to get insight into is how this risk depends on these three determining factors.

The technical details of the modeling and calculations can be found in Appendices A, B, and C.

## 3. Results

### 3.1. Epidemic Size in China

After calibration of the *SE*_2_ *I*_3_*R* model, we numerically calculated the final epidemic size (total cumulative number of cases) in China outside Hubei, using three different basic reproduction numbers and different control functions. The control functions were parametrized by *t*_∗_, which is the time after 23 January at which the control reaches its maximal value *u*_max_. Smaller *t*_∗_ corresponds to more rapid implementation of the control measures. In Figure 1, we plotted these cumulative numbers versus *t*_∗_, and we can observe that the epidemic final size is rather sensitive to the speed of implementation of the control measures. These curves also give upper bounds for the number of cumulative cases at any given time, assuming that the control efforts will be successful.

**Figure 1.**
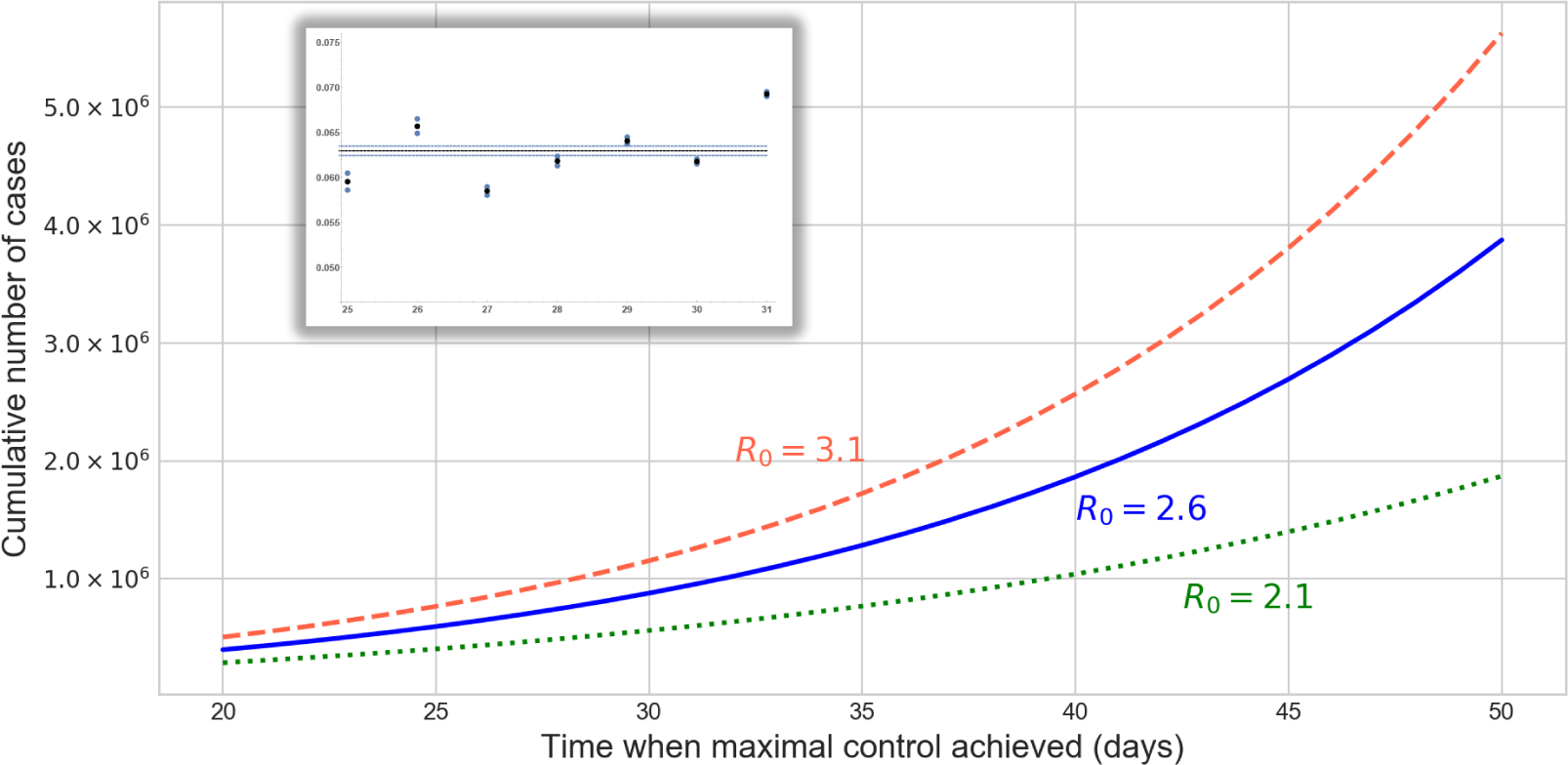
Final epidemic sizes in China, outside Hubei, with *R*_0_ = 2.1, 2.6, 3.1, as a function of the time when the control function *u*(*t*) reaches its maximum (in days after 23 January). Rapid implementation of the control generates much smaller case numbers. The inset shows the estimations of the ascertainment rate for the week 25–31, with average 0.063, based on the ratio of confirmed cases and the maximum likelihood estimates of the case numbers from exportation.

### 3.2. Risk of Major Outbreaks

We generated a number of plots to depict Risk(*C, θ, R*_loc_) for selected groups of countries from America, Europe, and Asia.

In the left of Figure 2, we can see the risks of American countries as functions of cumulative number of cases *C*, assuming each country has *R*_loc_ = 1.6 and their connectivity is their baseline *θ*. When *C* exceeds 600,000, with this local reproduction number and without any restriction in importation, outbreaks in the USA and Canada are very likely, while countries in South America (including Mexico), which are all in the green shaded region, still have moderate risks. To illustrate the impacts of control measures for the USA and Canada, we reduced *R*_loc_ to 1.4, and plotted the risks for different levels of reduction in connectivity to China, either due to travel restrictions or entry screening; see Figure 2 on the right. As the number of cases in China approaches one million, such reductions have a limited effect on the risk of outbreak. Figure 1 provides us with scenarios when *C* remains below certain values.

**Figure 2.**
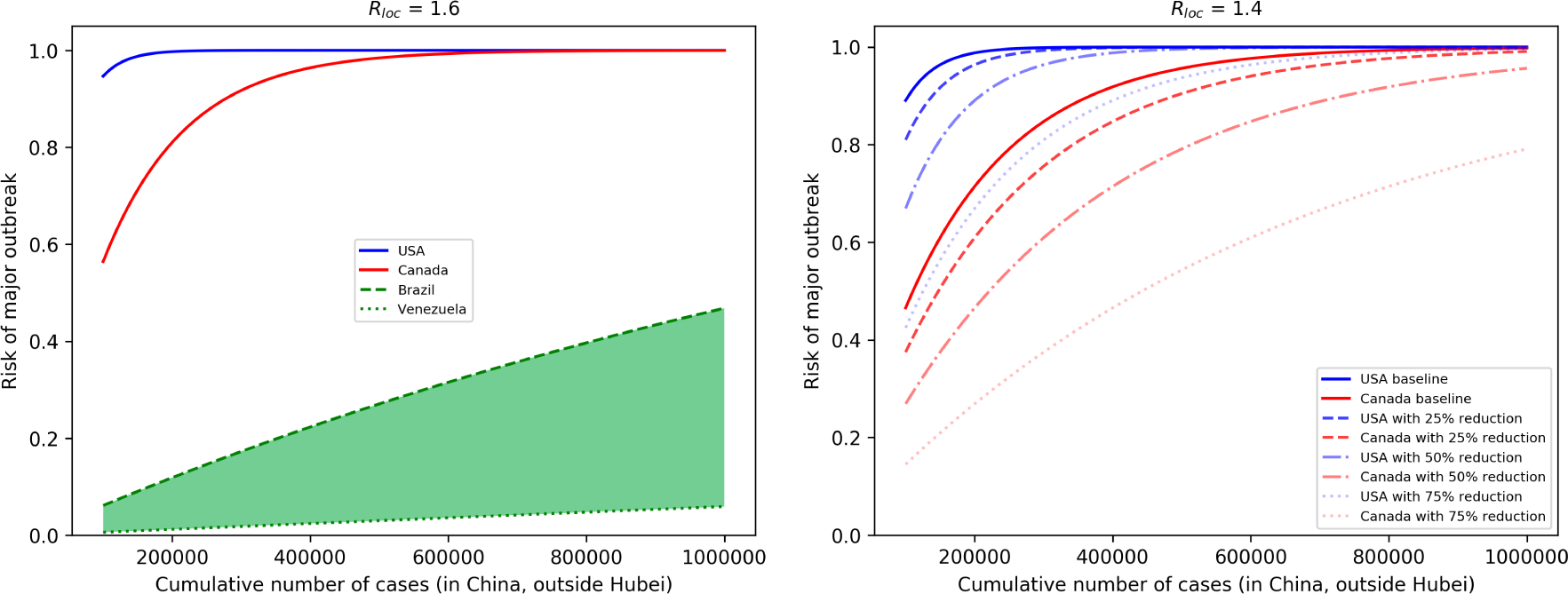
(**Left**) Risk of major outbreaks as a function of cumulative number of cases in selected countries, assuming *R*_loc_ = 1.6 and baseline connectivity to China. Other countries in South America, including Mexico, are inside the green shaded area. (**Right**) The effects of reductions of imported case numbers (either by travel restriction or entry screening) in the USA and Canada, assuming *R*_loc_ = 1.4.

We considered the group of countries from Asia which are the most connected to China: Thailand, Japan, Taiwan, and the Republic of Korea. They have similar baseline connectivity *θ*, and we focus on how travel restrictions and entry screenings can potentially reduce their risks, assuming different values of *R*_loc_ in the case *C* = 150,000 (on the left of Figure 3) and *C* = 600,000 (on the right of Figure 3). For illustration purposes, we plotted Thailand (red) and the Republic of Korea (blue), but Taiwan and Japan are always between those two curves. We can see that, for example, on the right of Figure 3 for *C* = 600,000, unless *R*_loc_ is very small, considerable reduction of the outbreak risk can be achieved only by extreme measures that prevent most importations.

**Figure 3.**
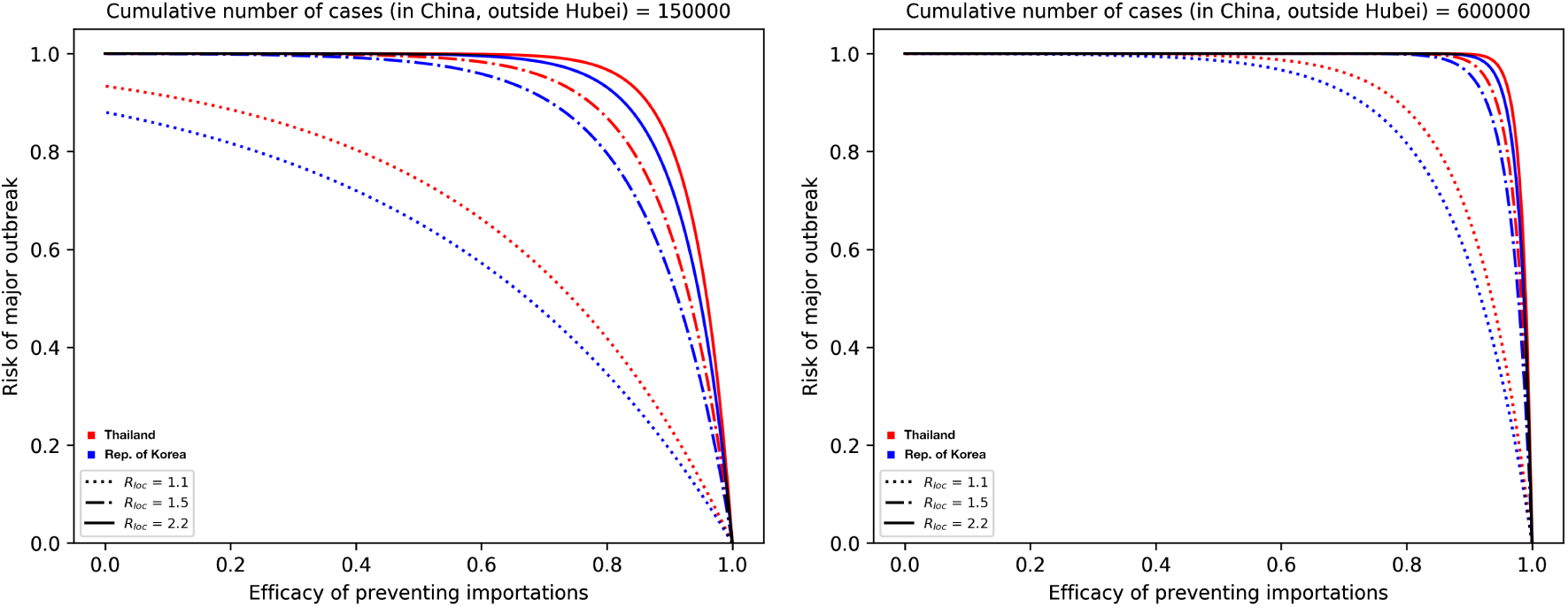
Outbreak risks for highly connected countries in Asia. Thailand and the Republic of Korea are plotted; the curves for Japan and Taiwan are in between them. (**Left**) We plot the risk vs. the efficacy of prevented importations when the cumulative number of cases reaches 150,000. (**Right**) *C* = 600,000. Black parts of the curves represent situations when the four countries are indistinguishable.

In Figure 4, we assumed that European countries have very similar *R*_loc_ and looked at their risks as a function of the number of cases. For illustration purposes, we selected countries which have relatively high (UK, Germany, France, Italy), medium (Belgium, Poland, Hungary), and low (Bulgaria, Croatia, Lithuania) connectivity to China. On the left, we assumed *R*_loc_ = 1.4 and baseline *θ*, and with these parameters, outbreaks will likely occur in high risk countries as the case number approaches one million. By reducing *R*_loc_ to 1.1 and by reducing *θ* to the half of its baseline (meaning that we assume that there is a 50% reduction in importations due to decreased travel and entry screenings), then the risk is significantly reduced, even with one million cases.

**Figure 4.**
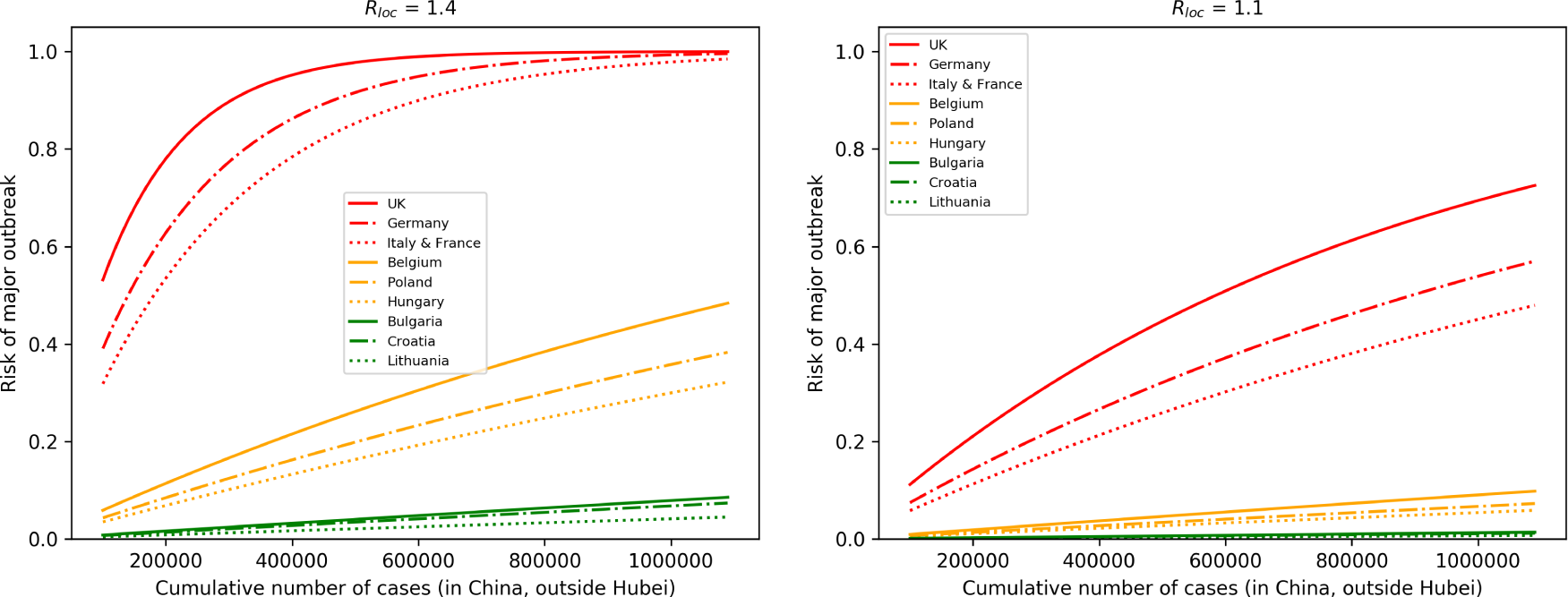
Selected European countries with high, medium, and low connectivity to China. (**Left**) The outbreak risk is plotted assuming their baseline connectivity *θ*, and *R*_loc_ = 1.4 for each country, as the cumulative number of cases is increasing. A significant reduction in the risks can be observed (**Right**), where we reduced *R*_loc_ to 1.1 and assumed a 50% reduction in importations.

### 3.3. Profile of Countries Benefiting the Most From Interventions

We also plotted the risks on a two-parameter map, as functions of *θ* and *R*_loc_. Observing the gradients of the risk map, we can conclude that countries with low connectivity but high *R*_loc_ should focus on further reducing importations by entry screening and travel restrictions, while countries with high connectivity but smaller *R*_loc_ better focus on control measures that potentially further reduce *R*_loc_. Countries in the middle benefit most from the combination of those two types of measures.

## 4. Discussion

By combining three different modelling approaches, we created a tool to assess the risk of 2019-nCoV outbreaks in countries outside of China. This risk depends on three key parameters: the cumulative number of cases in areas of China which are not closed, the connectivity between China and the destination country, and the local transmission potential of the virus. Quantifications of the outbreak risks and their dependencies on the key parameters were illustrated for selected groups of countries from America, Asia, and Europe, representing a variety of country profiles.

There are several limitations of our model, as each ingredient uses assumptions, which are detailed in the Appendices. There are great uncertainties in the epidemiological parameters as well. It is difficult to predict the epidemic trajectory in China, as the effects of the control measures are not clear yet. There were recent disruptions in international travel, suggesting that the EpiRisk parameters will not be accurate in the future. Nevertheless, when we have new information in the future about the case numbers in China, travel frequencies, efficacy of entry screenings, and local control measures, our method will still be useful for assessing outbreak risks.

We found that in countries with low connectivity to China but with relatively high *R*_loc_, the most beneficial control measure to reduce the risk of outbreaks is a further reduction in their importation number either by entry screening or travel restrictions (see Figure 5). Countries with high connectivity but low *R*_loc_ benefit the most from policies that further reduce *R*_loc_. Countries in the middle should consider a combination of such policies.

**Figure 5.**
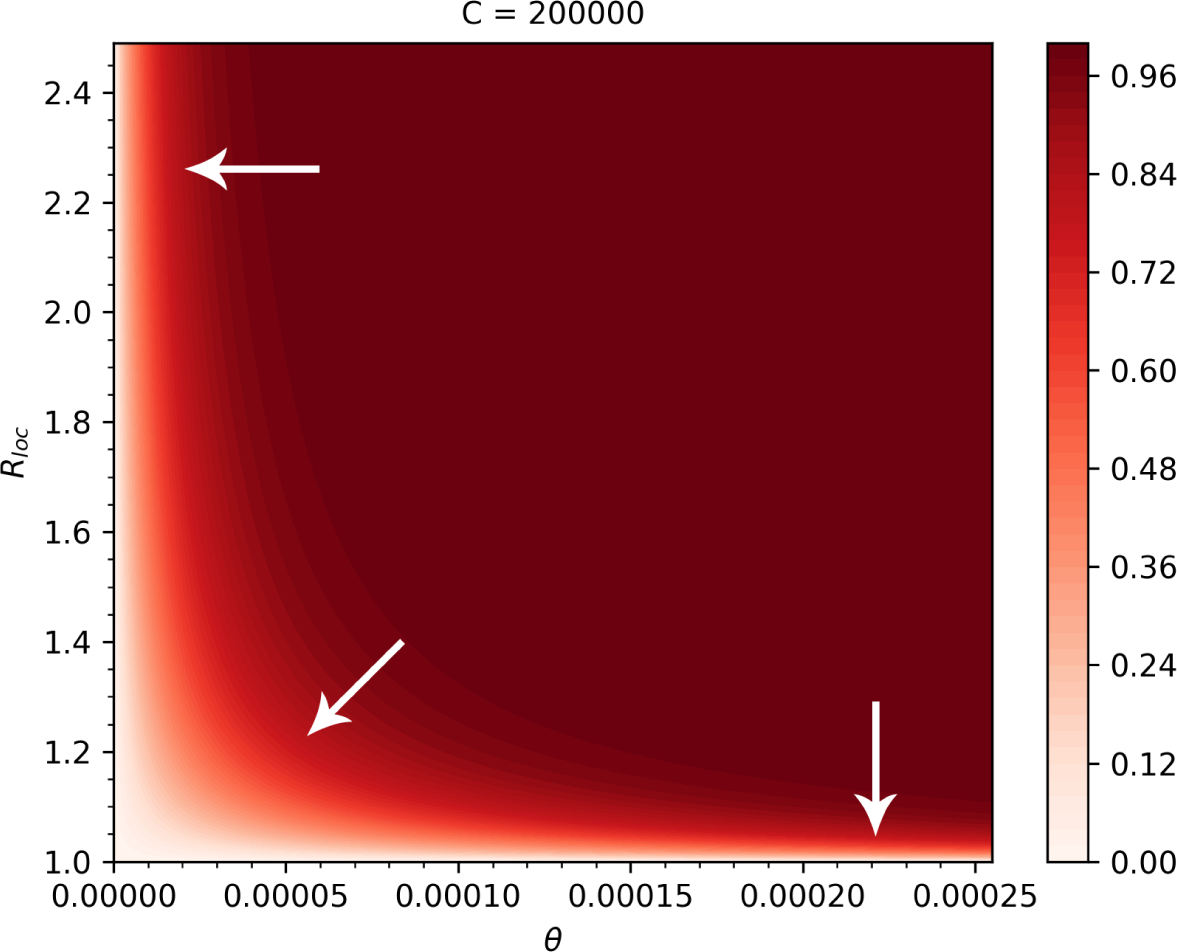
Heatmap of the outbreak risks as functions of *θ* and *R*_loc_, when *C* = 200,000. The arrows show the directions corresponding to the largest reductions in the risk.

Different control measures affect different key parameters. Several of these measures have been readily implemented in China, aiming to prevent transmissions. These are incorporated into our transmission model influencing the cumulative number of cases *C*. The connectivity *θ* may be affected, for example, by exit screening at Chinese airports, entry screening at the destination airport, and a decline in travel volume, all of which decrease the probability that a case from China will enter the population of the destination country. The parameter *R*_loc_ is determined by the characteristics and the control measures of the destination country. As new measures are implemented, or there is a change in travel patterns, these parameters may change in time as well.

Cumulative cases and connectivity can be estimated, in general. However, to make a good assessment of the outbreak risk, it is very important to estimate *R*_loc_ in each country. In the absence of available transmission data, one may rely on the experiences from previous outbreaks, such as the detailed description in [35] of the reductions in the effective reproduction numbers for SARS due to various control measures. In this study, we used a range of *R*_loc_ values between the critical value 1 and the baseline *R*_0_ = 2.6. A further source of uncertainty is in the distribution of the generation time interval, since a different distribution gives a different outbreak risk even with the same *R*_loc_. For our calculations, we used the distribution from [19] (see also [20]); a more in-depth discussion of this topic may be found in [36]. Knowing *R*_loc_ and the generation interval are needed not only to have a better quantitative risk estimation, but also for guidance as to which types of control measures may reduce the outbreak risk the most effectively.

## Data Availability

Links to all data used in the manuscript are provided.

http://www.epirisk.net

https://docs.google.com/spreadsheets/d/1wQVypefm946ch4XDp37uZ-wartW4V7ILdg-qYiDXUHM/htmlview?usp=sharing&sle=true

## Author Contributions

Conceptualization and methodology, G.R.; codes and computations A.D., T.T., F.A.B., P.B., G.R., and Z.V.; data collection and analysis, A.D., F.A.B., and T.T.; writing and editing, A.D., F.A.B., and G.R.; visualization, F.A.B., T.T., and Z.V. All authors have read and agreed to the published version of the manuscript.

## Appendix A Transmission Dynamics

The governing system of the transmission dynamics model is

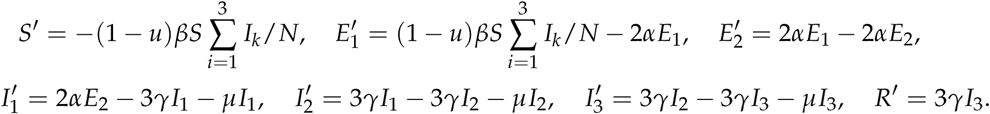

This is an extension of a standard SEIR model assuming gamma-distributed incubation and infectious periods, with the Erlang parameters *n* = 2, *m* = 3 (following the SARS-study [37]). Note that the choice of *n* = 2 is also consistent with the estimates summarized in Table 1. Given that disease fatalities do not have significant effect on the total population, we ignored them in the transmission model to ease the calculations (i.e., *µ* = 0 was used). In this model, the basic reproduction number is *R*_0_ = *β*/*γ*, the incubation period is *α*^−1^ and the infectious period is *γ*^−1^. The model is used to describe the disease dynamics in China outside Hubei province after 23 January. We assume that at time *t* after 23 January, an increasing control function *u*(*t*) represents the fraction of the transmissions that are prevented, thus the effective reproduction number becomes *R*(*t*) = (1 − *u*(*t*))*R*_0_*S*(*t*)/*N*.

Based on the previous estimates from the literature (see Table 1), we chose an incubation period *α*^−1^ = 5.1 days [30], basic reproduction number *R*_0_ = 2.6 (2.1–3.1) with the corresponding infectious period *γ*^−1^ = 3.3 (1.7–5.6) days [19]. To predict the final number of cases outside Hubei, we assume a gradually increasing control *u* from zero until a saturation point, and define *t*_∗_ the time when the eventual control *u*_max_ is achieved. The sooner this happens, the more successful the control is. Using the control term *u*(*t*) = min{*u*_max_*t*/*t*_∗_, *u*_max_}, disease control is reached at *t* = *t*_∗_(1 − 1/*R*_0_)/*u*_max_. For the calculations we choose *u*_max_ = 0.8, noting that such a drop in transmission has been observed for SARS, where the reproduction number was largely reduced by subsequent interventions [35]. With our baseline *R*_0_ = 2.6, disease control *R*(*t*) < 1 is achieved when *u*(*t*) > 0.615, meaning that more than 61.5% of potential transmissions are prevented, which occurs at time *t* = 0.77*t*_∗_.

Since the first case outside Hubei was reported on 19 January [5], for the initialization of the model we could assume that number of infected individuals on 23 January outside Hubei was equal to the number of cumulative cases outside Hubei up to that day. To calibrate the model, we estimated the number of cases from 24 January till 31 January outside Hubei based on case exportations, using the methodology of [15], assuming that exportations after 24 January were only from outside Hubei. Based on the maximal likelihood of case numbers that produce the observed number of exportations using EpiRisk [32], we estimate that the reported confirmed cases represent only 6.3% of the total cases for the regions outside Hubei (other estimates for ascertainment rate were: 5.1% [22], 10% in [38], and 9.2% (95% confidence interval: 5.0, 20.0) [39]), see the inset in Figure 1. The initial values for the exposed compartments in the SEIR model were selected such that the model output was consistent with the estimated case numbers outside Hubei between 24 January and 31 January. Solving the compartmental model, we obtained final epidemic sizes for various reproduction numbers and control efforts (see Figure 1), providing upper bounds for the cumulative number of cases *C* outside Hubei.

## Appendix B Calculating the Risk of Outbreaks by Importation

We create a probabilistic model to estimate the risk of a major outbreak in a destination country as a function of the cumulative number of cases *C* in China outside the closed areas, the local reproduction number *R*_loc_ in the destination country, and the connectivity *θ* between China and the destination country. We summarize these in Table A1.

**Table A1.** Parameters for calculating the risk of major outbreaks.

| Parameter | Interpretation | Depends on ... | Typical Range |
| --- | --- | --- | --- |
| $C$ | Cumulative case number in China, outside the closed areas | properties of nCoV-2019, efficacy of Chinese control | [100K, 6000K] |
| $R_{\text{loc}}$ | Local reproduction number in destination country | destination country | [1, 2.6] |
| $\theta$ | Probability of a importation chance that a case from the origin travelling to and mixing into the local population of the destination country | China and destination country | [0, 0.00025] |

We assume that the number of the imported cases entering the local population of the destination country follows a binomial distribution, i.e., the probability *p*_*i*_ corresponding to *i* imported cases in the destination country with connectivity *θ* to China is given by

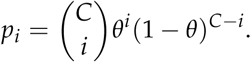

We calculate the extinction probability *z* of a branching process initiated by a new infection in the destination country. As in [19,20], we assume the number of secondary infections to follow a negative binomial distribution with generator function

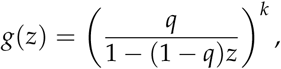

with dispersion parameter *k* and mean *µ* = *R*_loc_. Then, the probability parameter *q* of the distribution is obtained as 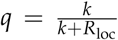. The extinction probability of a branch is the solution of the fixed point equation *z* = *g*(*z*).

Assuming that the destination country has *i* imported cases from China that are mixed into the local population, we estimate the probability of a major outbreak as the probability that not all the branches started by those *i* individuals die out, which is 1 − *z*^*i*^. Thus, the expectation of the risk of a major outbreak in country *x* can be calculated as

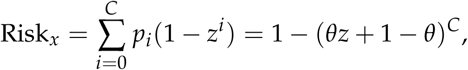

where we used the binomial theorem to simplify the sum. Having the input values of the parameters *C, θ, R*_loc_, with this model we can numerically calculate the risk.

## Appendix C

The codes for the computations were implemented in Mathematica and in Python, and they are available, including the used data, at [40].

